# Interaction of Post-Traumatic Stress Disorder and Race on Readmissions after Stroke

**DOI:** 10.1101/2023.07.26.23293224

**Authors:** Chen Lin, Peter H. King, Joshua Richman, Lori L. Davis

## Abstract

**Background:** There is limited research on outcomes of patients with post-traumatic stress disorder (PTSD) who also develop stroke, particularly regarding racial disparities. Our goal was to determine whether PTSD is associated with the risk of hospital readmission after stroke and if racial disparities existed.

**Methods:** The analytical sample consisted of all veterans receiving care in the Veterans Health Administration (VHA) who were identified as having a new stroke requiring inpatient treatment based on International Classification of Diseases codes. The retrospective cohort data was obtained from the VA Corporate Data Warehouse. The main outcome was any readmission to VHA. The hypothesis that PTSD is associated with readmission after stroke was tested using Cox regression adjusted for patient characteristics with PTSD as a time-varying covariate.

**Results:** Our final cohort consisted of 93,652 patients with inpatient stroke diagnosis and no prior VHA codes for stroke starting from 1999 with follow-up through 6-August-2022. Of these patients, 12,916 (13.8%) had comorbid PTSD. Of the final cohort, 16,896 patients (18.0%) with stroke were readmitted. Our fully-adjusted model for readmission found an interaction between African Americans (AA) and PTSD with a hazard ratio of 1.09 (95% CI 1.00-1.20; p<0.05). In stratified models, PTSD has a significant HR of 1.10 (1.02-1.18, p=0.01) for AA but not White veterans 1.05 (0.99-1.11, p=0.10).

**Conclusion:** Among AA Veterans who suffered stroke, pre-existing PTSD was associated with increased risk of readmission, which was not significant among White veterans. This study highlights the need to focus on high-risk groups to reduce readmissions after stroke.

## Introduction

Posttraumatic stress disorder (PTSD) has a lifetime prevalence of 6.1% in the U.S. adult population, ranging from 3.4% to 26.9% in civilian populations and 7.7% to 17.0% in military populations.^1,2 3, 4^ The economic burden on society is staggering – estimated at $232 billion annually in the U.S. for 2018.^5^ PTSD is an enduring disease that affects military veterans years after their service and for the remainder of their lives. Veterans of World War II and the Korean War had 12% prevalence rates 45 years after combat,^6^ 15% prevalence rates for Vietnam War veterans 15 years after combat,^4^ and 17% prevalence rates for military personnel returning from Iraq and Afghanistan.^7^ For this most recent group of veterans that served in Iraq and Afghanistan, PTSD is the most commonly diagnosed psychiatric disorder.^7^

PTSD has been identified as a major risk factor for developing various medical illnesses, including stroke.^8^ A meta-analysis found that the pooled hazard ratio showed that having PTSD was associated with a 59% higher risk of developing an incident stroke.^8^ The risk of developing cardiovascular disease and stroke is common to both civilian^9^ and veteran populations with PTSD.^10-12^ In a cohort of prisoners of war, those with severe and chronic stress had a higher risk of having a stroke.^13^ Additionally, young veterans less than 40 years old with PTSD have a 36% for increased risk of stroke compared with those veterans without PTSD.^14^

Unfortunately, there is limited research on outcomes of patients with PTSD who also develop stroke, particularly regarding racial disparities. In veterans with PTSD, worse treatment outcomes were seen for Black compared to White veterans.^15^ In patients with stroke, multiple areas of care are impacted by racial disparities, including those affecting outcomes such as recovery and readmission rates.^16, 17^ Readmission rates are an important outcome and priority area of interest for medical systems.^18^ Our main goal for this study was to determine whether patients diagnosed with PTSD who had a stroke have different risks for hospital readmission compared to patients who had a stroke without a PTSD diagnosis. Our secondary goal was to examine if race affected these rates. We hypothesized that patients with PTSD that had a stroke would be at higher risk of readmissions compared to those without PTSD.

## Methods

### Participants

The Research and Development Committee and the Institutional Review Board at the Birmingham Veterans Affairs Health Care System approved the study. The analytical sample consisted of all veterans receiving care in the Veterans Health Administration (VHA) who were identified as having an incident inpatient stroke without prior evidence of diagnostic codes for stroke in the VHA. Veterans could be included if they had an initial code for stroke from an outpatient encounter on the same day as their index inpatient hospitalization. Stroke diagnosis was identified by International Classification of Diseases (ICD) -9 codes 433.x1, 434.00, 434.x1, or 436.x or any ICD-10 code in the I63 group. The retrospective cohort data obtained for this study came from the VA Corporate Data Warehouse through the Veterans Administration Informatics and Computing Infrastructure (VINCI), which links all data from the VA electronic health record, death registry and outpatient pharmacy.

### Outcome

The main outcome was readmission to the VA with a diagnostic code for stroke coded as a time-to-event with date of last VHA encounter (inpatient or outpatient).

### Variables

Available covariates based on ICD codes included diabetes, hypertension, pulmonary hypertension, hyperlipidemia, myocardial infarction, congestive heart failure, valvular disease, peripheral vascular disease, drug abuse, alcohol abuse, and PTSD. Race and sex were obtained from VHA records based on self-report and age at incident stroke was normalized to have a mean of 0 and standard deviation of 1 so that model estimates are interpretable as the hazard ratio for a 1 standard deviation increase in age.

### Statistical Analysis

Descriptive statistics were calculated and differences between groups were tested using t-tests for continuous variables and chi-square tests for proportions. The study hypothesis that PTSD is associated with readmission after stroke with readmission related to stroke was explored using Kaplan-Meier curves and modeled using Cox regression adjusted for patient characteristics. While most comorbidities were present at the time of the index stroke, PTSD and other comorbidities were treated as time-dependent covariates to account for cases where new diagnoses emerged between the index stroke and stroke-related readmission. Pre-specified covariates of interest for the models were determined and we had a priori interest in racial disparities leading to the testing of the race interaction. Identification of PTSD, comorbidities, and other health conditions required at least two instances of appropriate ICD-9 or ICD-10 codes at least 30 days apart to avoid spurious diagnoses but were recorded as of the first date, a method used for other VHA datasets.^19^ Examination of stratified Kaplan-Meier curves suggested a possibly meaningful interaction between race and PTSD. After formally testing for an interaction in minimally adjusted and fully adjusted Cox models, we presented adjusted model results stratified by race.

## Results

Our final cohort consisted of 93,652 patients with inpatient stroke diagnosis and no prior VHA codes for stroke from 1999 identified up to 20-April-2022 with follow-up for readmission through 6-August-2022. Of these patients, 12,916 (13.8%) also had comorbid PTSD at the time of their stroke. Of the final cohort, 16,896 patients (18.0%) with an index diagnosis of stroke were readmitted for any cause to a VA hospital facility. Table 1 describes readmissions information, length of follow-up, and medical comorbidities of our population, stratified by race. In our population, 97.1% were males, 62.89% were White, and 22.4% were African-American. Common comorbidities included: Type 2 Diabetes Mellitus (67.3%), hyperlipidemia (72.8%), and hypertension (92.9%). Table 2 describes readmissions information and medical comorbidities stratified by race and pre-existing diagnosis of PTSD prior to stroke. We noted evidence of an interaction between race and PTSD as risks for stroke-related readmission, with a p-value of 0.02 in a model adjusted only for race and PTSD (with interaction) and p=0.06 in a fully-adjusted model.

**Table 1.**
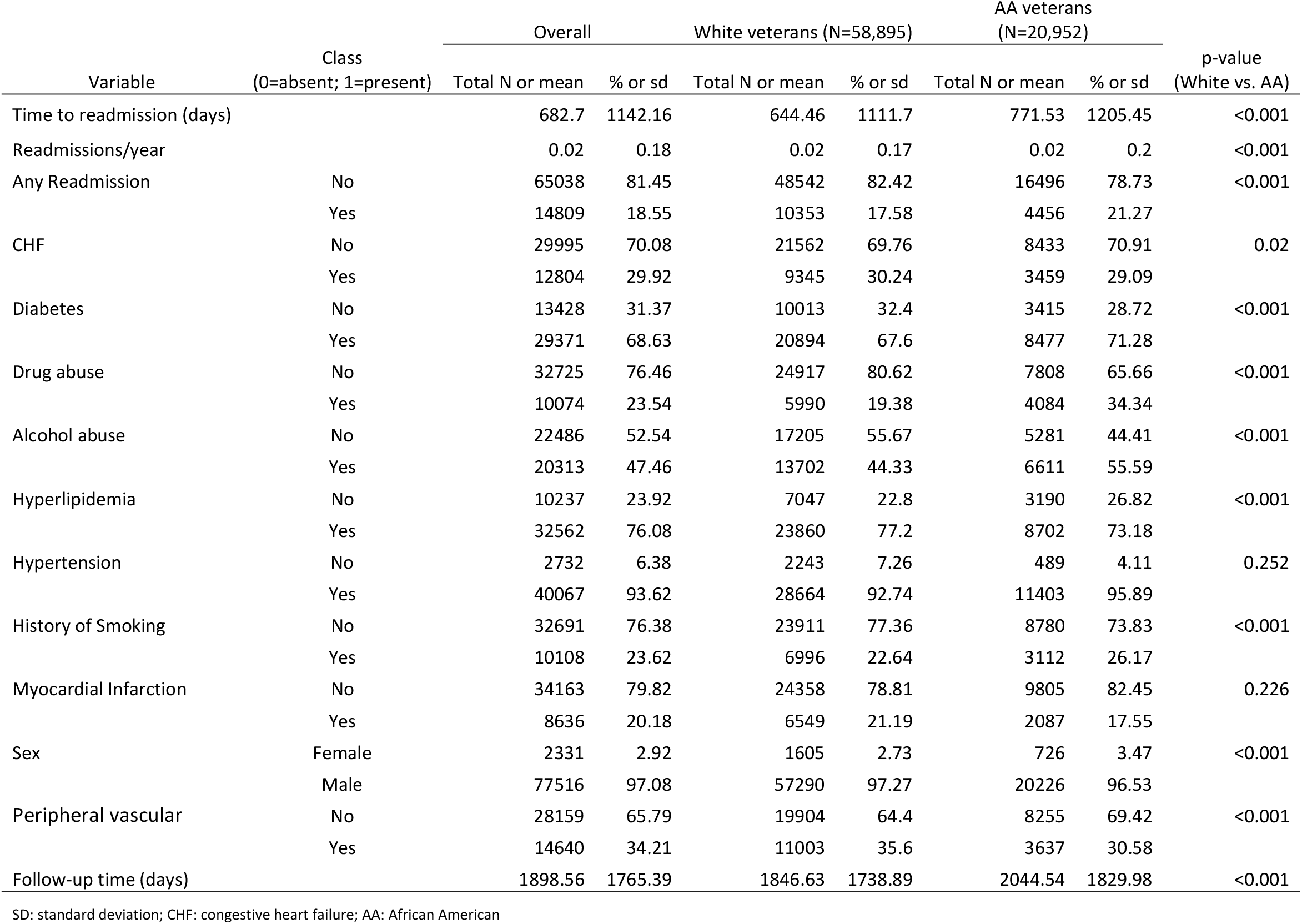
Characteristics of patients with readmission after stroke by race.

**Table 2.**
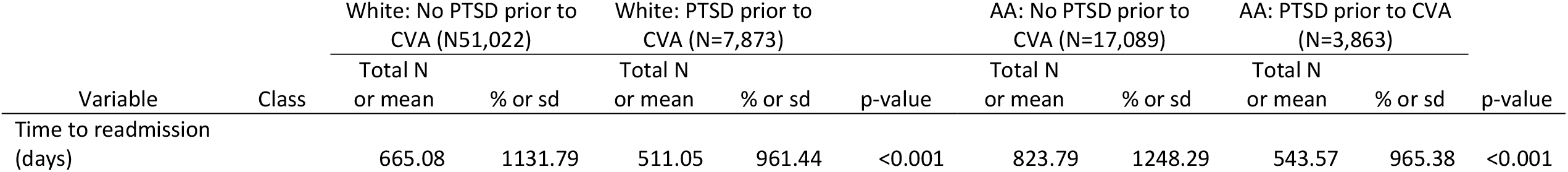
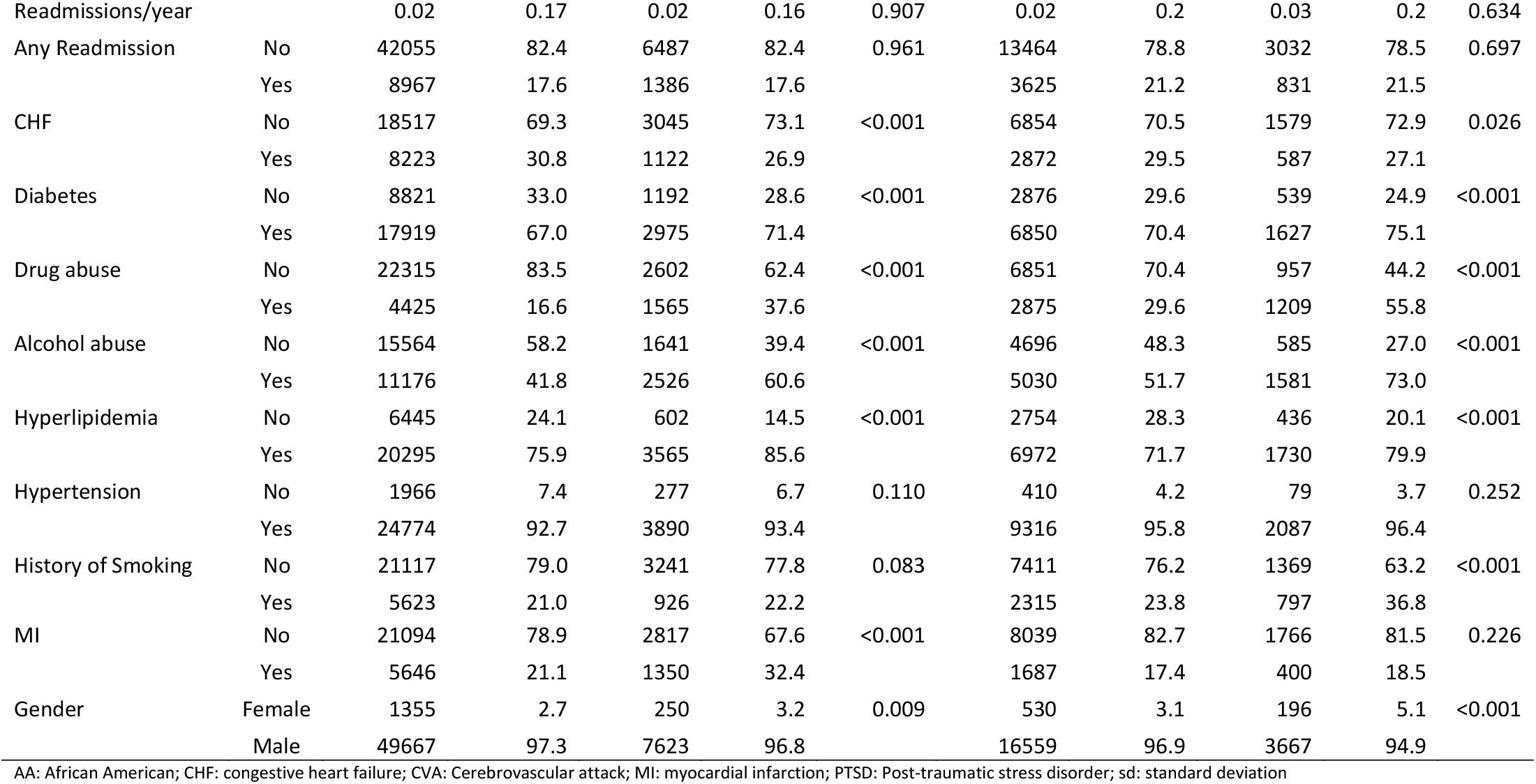
Characteristics of patients with readmission after stroke by race and pre-existing PTSD.

The probability of not having a readmission after stroke over time is shown in Figure 1, plotted separately by race (White and African American) with PTSD treated as a time-dependent covariate. At cohort entry, 13.4% of White veterans had PTSD, compared to 18.4% of African American veterans. Both White and African American veterans with PTSD had higher rates of type 2 diabetes mellitus, substance abuse including illegal drugs and alcohol, hyperlipidemia, and smoking compared to their respective groups without PTSD. Both White and African American veterans without PTSD had higher rates of congestive heart failure compared to their respective groups with PTSD. Both groups had similar rates of hypertension. On average for African American veterans, 0.02% were readmitted per year of follow-up for those without PTSD and 0.03% for those with PTSD. On average for White veterans, 0.02% were readmitted per year of follow-up in both those with and without PTSD.

**Figure 1.**
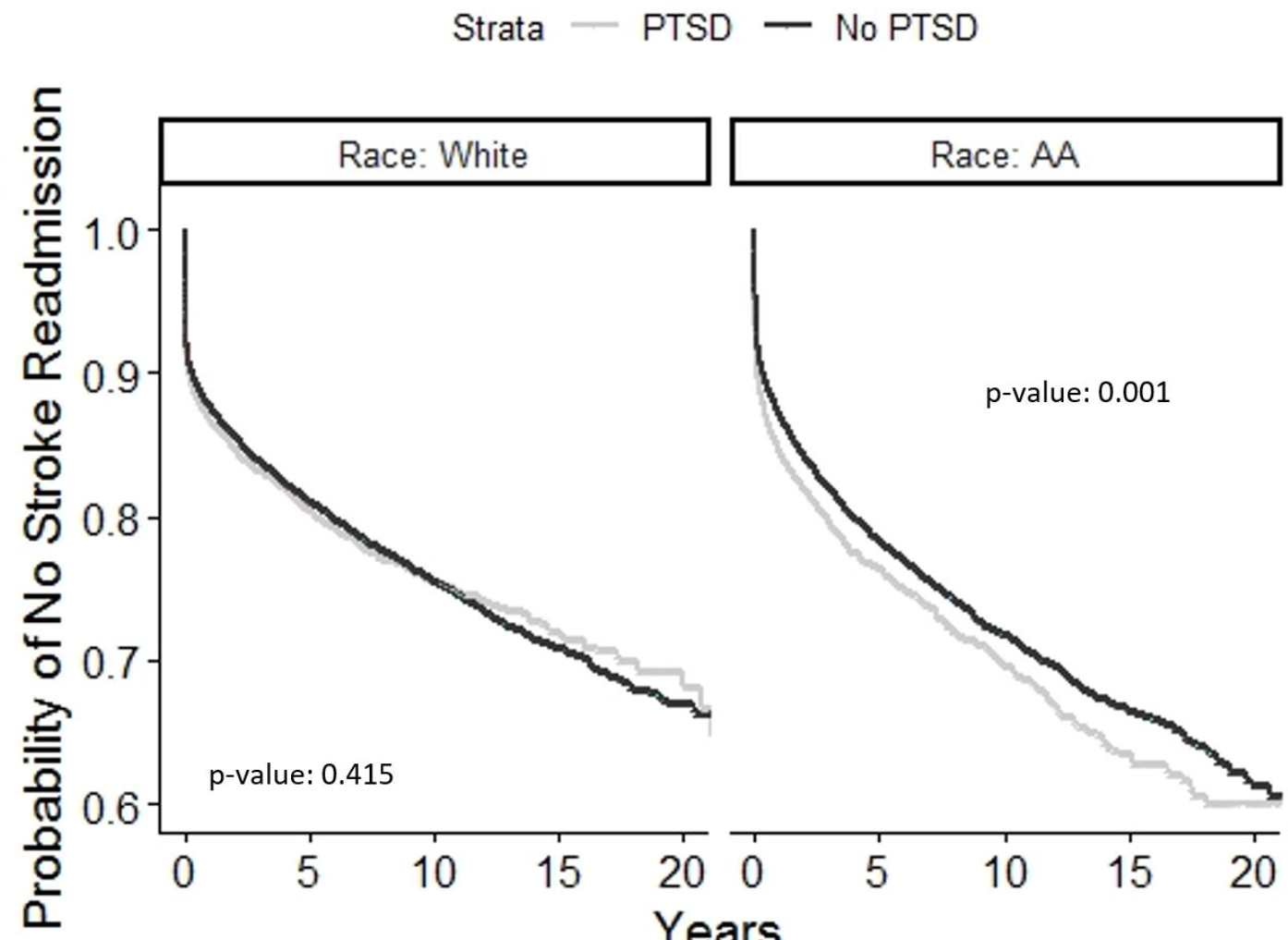
Stroke Readmission by Race and PTSD PTSD: Post-traumatic stress disorder; AA: African-American

Our fully adjusted model for readmission, both overall and stratified by race, is presented in Table 3. Our fully-adjusted model found African Americans with PTSD who had a stroke had a hazard ratio of 1.09 (95% CI 1.00-1.20; p<0.05) for readmissions in the overall veteran population. In the overall population, other significant variables that increased the risk for readmission included: age (HR 1.01, 95% CI 1.00-1.01; p=<0.01), congestive heart failure (HR 1.11, 95% CI 1.05-1.18; p=<0.01), type 2 diabetes (HR 1.06, 95% CI 1.01-1.11; p=0.01), history of myocardial infarction (HR 1.17, 95% CI 1.10-1.25; p=<0.01), and peripheral vascular disease (HR 1.16, 95% CI 1.11-1.23; p=<0.01). Variables that decreased risk included: hypertension (HR 0.87, 95% CI 0.83-0.91; p=<0.01) and hyperlipidemia (HR 0.93, 95% CI 0.89-0.98; p=<0.01).

**Table 3.**
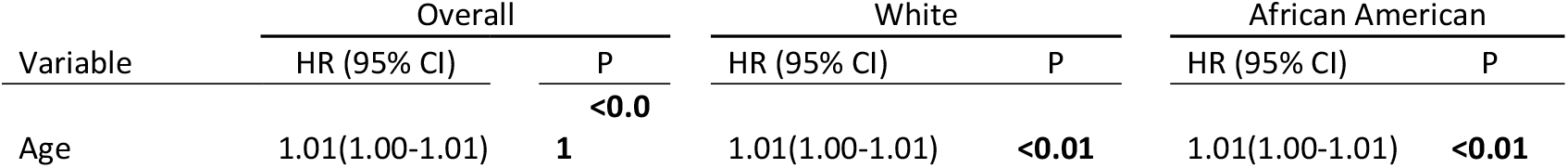
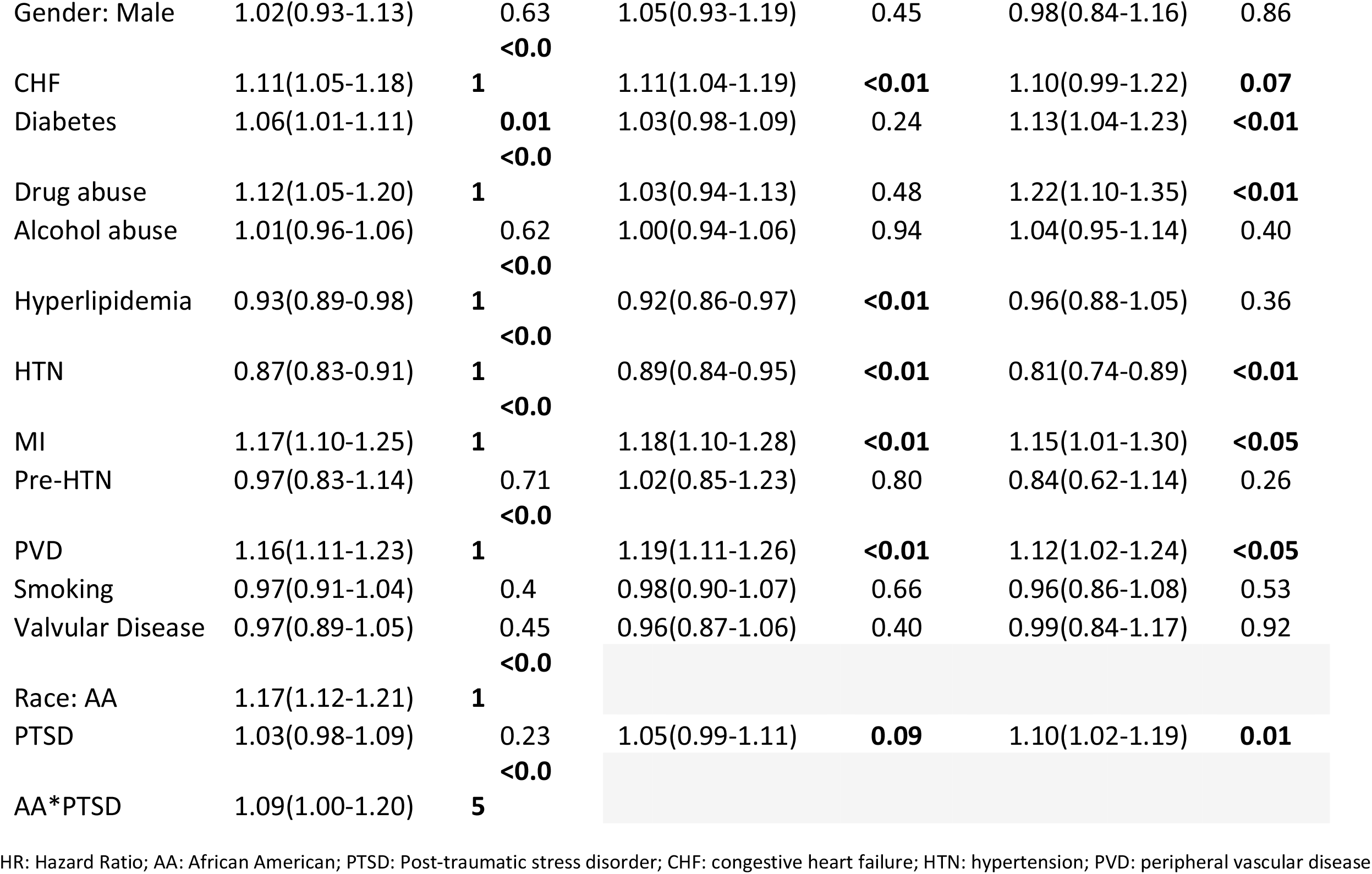
Adjusted model for readmission in patients after stroke.

Within the African American veteran population that had stroke, those with PTSD had a hazard ratio of 1.10 (95% CI 1.02-1.19; p=0.01) for readmissions shown in Table 2. Other significant variables that increased the risk for readmissions included: age (HR 1.01, 95% CI 1.00-1.01; p=<0.01), type 2 diabetes (HR 1.13, 95% CI 1.04-1.23; p<0.01), drug abuse (HR 1.22, 95% CI 1.10-1.35; p<0.01), history of myocardial infarction (HR 1.15, 95% CI 1.01-1.30; p=0.03), and peripheral vascular disease (HR 1.12, 95% CI 1.02-1.24; p=0.02). Within the White veteran population, those that had PTSD had a hazard ratio of 1.05 (95% CI 0.99-1.11; p=0.09) for readmissions. Other significant variables that increased the risk for readmissions included: age (HR 1.01, 95% CI 1.00-1.01; p=<0.01), congestive heart failure (HR 1.11, 95% CI 1.04-1.19; p=<0.01), history of myocardial infarction (HR 1.18, 95% CI 1.10-1.28; p=<0.01), and peripheral vascular disease (HR 1.19, 95% CI 1.11-1.26; p=<0.01). Hypertension decreased the risk for readmissions in both African American (HR 0.81, 95% CI 0.74-0.89; p=<0.01) and White veterans (HR 0.89, 95% CI 0.84-0.95; p=<0.01). Hyperlipidemia was associated with a lower risk only in White veterans (HR 0.92, 95% CI 0.86-0.97; p=<0.01); whereas only in African Americans were type 2 diabetes (HR 1.13, 95% CI 1.04-1.23; p=<0.01) and drug abuse (HR 1.22, 95% CI 1.10-1.35; p=<0.01) associated with an increased risk for readmission.

## Discussion

In a large cohort of veterans who have been diagnosed with a stroke, we found that those with pre-existing PTSD had an increased risk of all-cause readmissions than those without PTSD. Additionally, we found that African Americans with PTSD who suffered stroke were more likely than their counterparts without PTSD to be re-hospitalized. This is one of the first studies showing that patients who have stroke and suffer from pre-existing PTSD have worse outcomes as suggested by a higher risk for re-hospitalization. This study underscores the importance of developing strategies to prevent re-hospitalization for this at-risk population.

PTSD has been associated with a significant increased risk of developing an ischemic stroke as an independent risk factor.^12, 14^ Separately, patients without pre-existing PTSD have also been shown to be at risk of developing post-stroke PTSD.^20-22^ Patients that develop post-stroke PTSD have also been shown to have worse outcome.^20, 22, 23^ In survey studies of patients that develop post-stroke PTSD, patients have had worse quality of life and more issues with medication adherence.^22, 23^ In our cohort, veterans with PTSD had higher rates of cardiac conditions and smoking, all of which likely contributed to readmission. A higher risk for readmission in African American veterans was associated with drug abuse and type 2 diabetes. A logical focus in the development of strategies to prevent re-admissions for these veterans would target these high-risk comorbidities. Our results help bridge the gap in knowledge of post-stroke outcomes in patients with PTSD showing that these patients are at higher-risk for being re-admitted. Prospective studies monitoring this high-risk population are needed to identify safety net measures that may reduce readmissions.

Reducing readmissions is a key priority for medical systems.^18^ While this study did not examine reasons for readmission, we did follow patients for 20 years after their initial stroke admission. Studies on post-stroke readmissions have generally focused on identifying predictors based on medical comorbidities.^24, 25^ Most studies examine how post-stroke mental health issues lead to worse outcomes, but few focus on readmissions.^26-28^ Stein et al.^19^ examined whether depression after ischemic stroke and myocardial infarction led to more readmissions in the Nationwide Readmissions Database. They found that not only was depression after stroke more likely to lead to readmission, but that the risk was greater than with depression after myocardial infarction.^26^ In our veteran cohort, veterans with PTSD had increased readmissions compared to those without PTSD. A racial disparity was also seen with African American having higher risk for readmission compared to White veterans.

Our finding that African Americans veterans who suffer stroke had higher risk of readmission if previously diagnosed with PTSD parallels findings in other populations. In the civilian population, African Americans tended to have higher rates of readmission.^29^ For veterans, one recent study examining hospital readmissions following hospitalization for COVID-19 also found higher rates of readmission in African American veterans. Interestingly, in our study, White and African Americans with PTSD shared many similar medical comorbidities (hypertension, smoking, etc.) compared to their counterparts without PTSD, suggesting that other factors are at play for the higher risk of readmissions. There are likely issues not covered by our analysis including socioeconomic status, caregiver support, and community support that may contribute to the higher risk of all-cause readmissions in the African American veteran population. We did see in our fully adjusted model that type 2 diabetes and drug abuse were associated with a higher readmission risk, but not in White veterans. Our study is the first to find that African Americans veterans with stroke are at higher risk of all-cause readmissions than non-African Americans, particularly if they also suffer from PTSD.

Recent trends in declining readmissions nationwide have been tied to hospitals’ responding to incentives to reduce readmissions.^18^ The VHA has been collaborating with the Centers for Medicare and Medicaid Services to report hospital performance and reduce all-cause readmission rates.^30^ Within the VHA, readmission rates for patients with heart failure have been declining.^31^ However, among those veterans admitted with cardiac issues or pneumonia, those admitted at VHA hospitals were associated with higher 30-day risk-standardized all-cause readmission rates than those admitted to non-VHA hospitals, both nationally and within similar geographic areas.^30^ In patients with stroke, a Centers for Medicare & Medicaid Services and VHA retrospective study compared a basic Medicare and Medicaid model to a more comprehensive model. The comprehensive model included focusing on social factors such as homelessness and clinical factors such as stroke severity. However, neither model predicted readmission rates.^32^ While this study identified those with a mental health disorder, neither a PTSD-specific diagnosis nor symptoms were recorded.

Our results highlight an important area for potential improvement in future programs to assess and prevent readmissions for veterans with stroke both with and without PTSD in VHA hospitals. So far, the data available have not accounted for the impact of PTSD symptoms which are more frequent in the veteran population. An improved way to account for PTSD symptoms and link them with readmission data is necessary. Additionally, incentives have been shown to improve readmission rates.^18^ Future interventions to reduce readmission rates in patients with stroke should also consider monitoring and treating PTSD symptoms. Our findings indicate that post-discharge transitional care after stroke and monitoring for PTSD symptoms after stroke could be particularly important in African American veterans.

## Limitations

The present study has several strengths, including its large cohort but also has several limitations.

First, because this is a retrospective national cohort study, it is subject to inherent biases, such as residual confounding. Second, we used ICD codes to identify patients rather than primary clinical data. Although we included patients with the diagnosis of stroke, data regarding type of stroke including ischemic or hemorrhagic were not distinguished. Third, women were under-represented in our study and limits the generalizability. Fourth, because an administrative dataset was used, this data often has errors in diagnosis code allocation. Finally, because the VHA database only captures VHA readmissions, other hospital readmissions were not captured unless they were also transferred to a VHA hospital. This means we likely underestimated readmissions rates based on conservative estimates of readmission.

## Conclusion

Among AA Veterans who suffered stroke, PTSD was associated with increased readmission, but the association was not significant among White veterans. Certain risk factors, such as diabetes and drug abuse, were also associated with increased risk for readmission in AA but not White veterans. This study highlights the need for further investigation into this high-risk group including the development of post-discharge transitional care plans to reduce readmissions after stroke.

## Data Availability

Data are available from the authors upon reasonable request and with permission from the Veterans Affairs Research Offices.

## Study Funding

Dr. Lin is supported by VA IK2 CX002104 and VA I21 RX003612.

## Notes

**Disclosure of Conflicts of Interest:** The authors have no conflicts of interest.

### Competing Interest Statement

The authors have declared no competing interest.

### Clinical Trial

N/A

### Author Declarations

Birmingham VA IRB approved the study.

